# Subthalamic deep brain stimulation alleviates motor symptoms without restoring deficits in corticospinal suppression during movement preparation in Parkinson’s disease

**DOI:** 10.1101/2023.08.03.23293486

**Authors:** Emmanuelle Wilhelm, Gerard Derosiere, Caroline Quoilin, Inci Cakiroglu, Susana Paço, Christian Raftopoulos, Bart Nuttin, Julie Duque

**Affiliations:** Institute of Neuroscience, Catholic University of Louvain, 1200 Brussels, Belgium; Department of Adult Neurology, Saint-Luc University Hospital, 1200 Brussels, Belgium; NOVA IMS, Universidade Nova de Lisboa, 1070-312 Lisbon, Portugal; Department of Neurosurgery, Saint-Luc University Hospital, 1200 Brussels, Belgium; Department of Neurosurgery, UZ Leuven, 3000 Leuven, Belgium

**Keywords:** Parkinson’s disease, subthalamic nucleus, deep brain stimulation, transcranial magnetic stimulation, motor control

## Abstract

**Background:** Parkinson’s disease (PD) patients exhibit alterations in neurophysiological mechanisms underlying movement preparation, especially the suppression of corticospinal excitability – called “preparatory suppression” – considered to propel movement execution by increasing motor neural gain in healthy individuals.

**Objective:** Deep brain stimulation (DBS) of the subthalamic nucleus (STN) being an attractive treatment for advanced PD, we aimed to investigate the potential contribution of this nucleus to PD-related changes in such corticospinal dynamics.

**Methods:** On two consecutive days, we applied single-pulse transcranial magnetic stimulation over both primary motor cortices in 20 PD patients treated with bilateral STN-DBS (ON vs. OFF), as well as 20 healthy control subjects. Motor-evoked potentials were elicited at rest or during a left- or right-hand response preparation in an instructed-delay choice reaction time task. Preparatory suppression was assessed by expressing amplitudes of motor potentials evoked during movement preparation relative to rest.

**Results:** Advanced PD patients exhibited a deficit in corticospinal suppression during movement preparation, limited to the responding hand (especially the most-affected), independently of STN-DBS. Significant links between preparatory suppression and clinical variables were found for least-affected hands only.

**Conclusion:** Our study provides evidence of altered corticospinal dynamics during movement preparation in advanced PD patients treated with STN-DBS. Consistent with results in earlier-stage patients, preparatory suppression deficits were limited to the responding hand and most pronounced on the most-affected side. STN-DBS did not restore this abnormality, which warrants further investigations into possible neuroanatomical sources of such corticospinal suppression, necessary to understand the consistent lack of this mechanism in PD patients.

## Introduction

Dysfunctional cortico-basal ganglia (BG)-thalamocortical circuits, following nigrostriatal dopaminergic neurodegeneration, are an integral part of Parkinson’s disease (PD).^1, 2^ Targeting core structures within these circuits with deep brain stimulation (DBS) has become an increasingly attractive treatment for advanced PD,^3^ as it represents an efficient second-line option once clinical benefits of oral dopamine replacement therapy (DRT) fade.^4^ The main target for DBS in PD is the subthalamic nucleus (STN),^5–7^ a key region within the BG involved in human motor control.^8–10^

The exact mechanisms underlying clinical benefits of STN-DBS in PD are complex and not fully elucidated yet:^3, 11, 12^ dysfunctional oscillatory activity involving the STN appears to be disrupted with DBS, ^13–16^ preventing excessive movement inhibition and thus alleviating motor symptoms.^17–19^ Yet, in some patients, STN-DBS can generate side effects, such as altered inhibitory control resulting in more impulsive behaviour.^20–23^

Therefore, based on the need to optimize STN-DBS and minimize side effects, the interest in understanding functions and neural dynamics of this nucleus has been growing steadily.^8^ Within the field of motor control, its role has been largely examined in the context of action stopping,^24–28^ including recent studies on alterations in corticospinal excitability (CSE) during non-selective motor inhibition in patients with STN-DBS.^29^ However, the potential contribution of the STN to PD-related changes in dynamics of CSE during action preparation has not been explored,^9, 30^ and represents the aim of the current study.

Interestingly, when applying single-pulse transcranial magnetic stimulation (TMS) over the primary motor cortex (M1) of healthy subjects, a transient drop in motor-evoked potential (MEP) amplitudes has been observed consistently during action preparation, indicating a paradoxical suppression of CSE preceding movement execution.^30–33^ Among other hypotheses, this so-called “preparatory suppression” has been considered to facilitate movement release and speed by increasing motor neural gain.^33–37^ Accordingly, in healthy individuals, the strength of preparatory suppression correlates with the speed of motor responses.^33, 34^ Importantly, we could recently uncover first deficits of such corticospinal suppression in PD patients, which were consistent with PD dominance and evolved with disease duration and levels of motor slowness in the same hand.^38^

In the present study, our goal was to investigate such markers of intact movement preparation in advanced PD patients treated with STN-DBS. In line with our previous findings, we expected a lack of preparatory suppression, especially in the most-affected hand, in the absence of stimulation. Since DBS alleviates motor symptoms even at advanced disease stages, we wanted to test the hypothesis that ON-DBS, patients would exhibit a less pronounced deficit of this mechanism.

To address these hypotheses, in two sessions on two consecutive days, we applied TMS over M1 in PD patients implanted with bilateral STN-DBS while they executed an instructed-delay choice reaction time (RT) task. Even though TMS has been safely performed in DBS patients in previous studies,^39–41^ CSE investigations in DBS patients are remarkably rare, especially with concurrent behavioural measures.^29^ Here, patients were tested either ON- or OFF-DBS, while taking their regular DRT in both sessions; they were matched with healthy controls (HCs), also performing two sessions on consecutive days. Neurophysiological measures were cross-analysed with demographic and clinical data.

## Methods

We recruited 22 patients with advanced idiopathic PD; two were excluded from the analysis, since they were unable to execute the task OFF-DBS.^42^ The remaining patients (6 females, 62.4 ± 7.9 years) were matched with 20 HC subjects, for age (62.4 ± 8.0 years), gender and education. All controls and 15 patients were right-handed.^43^ Exclusion criteria comprised presence of severe cognitive decline (based on Montreal Cognitive Assessment)^44, 45^ and history of major psychiatric, neurological (except PD) or substance use disorder (except nicotine). Participants gave written informed consent, following a protocol approved by the Biomedical Ethics Committee of Saint-Luc University Hospital (Brussels, Belgium), in accordance with the Declaration of Helsinki.

Demographic and clinical features of patients are presented in Table 1; they were diagnosed based on clinical criteria by the Movement Disorder Society (MDS)^46–48^ and recruited through the Neurology and Neurosurgery departments of Saint-Luc and UZ Leuven university hospitals (Belgium). All patients had undergone bilateral electrode implantation at least six months before participating in this study; all were taking DRT and physically independent enough to visit our laboratory. None was tremor-dominant nor displayed impulse control disorders, dopamine dysregulation syndrome or dyskinesias around the time of the experiment.^49^

**Table 1.** Details of demographic and clinical data in PD patients.

| Patient | Gender | Handedness | Disease duration (years) | Time since surgery (months) | Hoehn & Yahr | MDS-UPDRS-III, Total, OFF-DBS | MDS-UPDRS-III, Total, ON-DBS | LEDD (mg) | PD dominance (MA side based on MDS-UPDRS-III) | MoCA | BDI-II | UPPS |
| --- | --- | --- | --- | --- | --- | --- | --- | --- | --- | --- | --- | --- |
| 1 | M | L | 7 | 11 | 2 | 27 | 18 | 600 | R | 29 | 3 | 83 |
| 2 | M | R | 8 | 6 | 2 | 39 | 18 | 1911 | R | 26 | 17 | 93 |
| 3 | M | R | 8 | 8 | 2 | 25 | 16 | 980 | L | 29 | 0 | 79 |
| 4 | M | L | 8 | 12 | 3 | 61 | 31 | 604 | R | 28 | 22 | 79 |
| 5 | M | L | 10 | 24 | 2 | 19 | 14 | 800 | R | 29 | 28 | 99 |
| 6 | M | R | 10 | 7 | 2 | 42 | 43 | 505 | R | 25 | 0 | 94 |
| 7 | M | R | 11 | 17 | 2 | 42 | 30 | 1083 | R | 29 | 18 | 110 |
| 8 | F | R | 12 | 8 | 2 | 27 | 20 | 637 | L | 27 | 6 | 93 |
| 9 | M | R | 13 | 16 | 2 | 35 | 26 | 960 | R | 26 | 0 | 113 |
| 10 | M | L | 14 | 48 | 2 | 47 | 25 | 525 | L | 29 | 14 | 97 |
| 11 | M | R | 15 | 18 | 2 | 16 | 14 | 710 | R | 28 | 14 | 92 |
| 12 | M | R | 15 | 24 | 2 | 43 | 34 | 800 | L | 28 | 9 | 73 |
| 13 | M | L | 15 | 53 | 2 | 46 | 22 | 50 | L | 28 | 6 | 70 |
| 14 | F | R | 16 | 24 | 2 | 44 | 35 | 883 | L | 29 | 28 | 93 |
| 15 | F | R | 16 | 120 | 3 | 57 | 38 | 2075 | L | 28 | 27 | 104 |
| 16 | M | R | 18 | 8 | 2 | 30 | 20 | 776 | L | 25 | 10 | 100 |
| 17 | M | R | 18 | 8 | 3 | 44 | 28 | 465 | L | 21 | 16 | 90 |
| 18 | F | R | 18 | 6 | 2 | 24 | 11 | 610 | L | 30 | 7 | 89 |
| 19 | F | R | 20 | 60 | 4 | 78 | 70 | 605 | L | 26 | 41 | 111 |
| 20 | F | R | 20 | 16 | 3 | 64 | 47 | 742 | L | 25 | 23 | 106 |
| Mean/% | 70% M | 75% R | 163.2 | 24.7 | 2.3 | 40.5 | 28 | 816.1 | 40% R | 27.3 | 14.5 | 93.4 |
| SD | / | / | 50.1 | 27.5 | 0.6 | 16.0 | 14.1 | 460.5 | / | 2.2 | 11.2 | 12.3 |

Patients are ranged according to their disease duration, that is, the time since their diagnosis (in years). Data from outlier patient 8 are shown in grey. Abbreviations: PD = Parkinson's disease; MDS-UPDRS-III = Movement Disorder Society Unified Parkinson's Disease Rating Scale, Part III; DBS = deep brain stimulation; ON-DBS = session performed with patients having taken their usual PD medication and with their DBS device kept on; OFF-DBS = session performed with patients having taken their usual PD medication, but after a wash-out period of 30-45 minutes after having turned off their DBS device; LEDD = Levodopa Equivalent Daily Dose; MA = Most-affected; MoCA = Montreal Cognitive Assessment; BDI-II = Beck Depression Inventory, Second Edition; UPPS = UPPS Impulsive Behaviour Scale; NA = not available; SD = Standard deviation.

Experiments were performed at the Institute of Neuroscience of *UCLouvain* (Brussels, Belgium). Every participant underwent two testing sessions at the same hour on two consecutive mornings. Patients performed one session with their stimulator on (“ON-DBS”) and another with their stimulator turned off (“OFF-DBS”), in a randomised order. The programming tablet was connected on both days and neither the patient nor the TMS experimenter was informed of the session order; patients were asked not to reveal the DBS condition in case they sensed it (which regularly happened). In both sessions, motor scoring, was performed after a “washout” period of 30 minutes^17, 23, 50^ (in order not to reveal the DBS condition), using the MDS-Unified Parkinson’s Disease Rating Scale Part III (MDS-UPDRS-III). The latter served to obtain a global score of motor impairment and a bradykinesia subscore (see Supplementary Material). Since all patients still took DRT, for both sessions, data acquisition was performed within the same delay with respect to their last medication dose;^29^ note, however, that in our previous study on preparatory suppression in PD, DRT did not impact the latter.^38^ Matched HCs performed two identical sessions on two consecutive mornings, turned into fictive OFF- and ON-DBS sessions. Both groups of participants filled in the French version of two self-evaluation questionnaires, validated for the use in PD:^51–53^ the Beck Depression Inventory Second Edition (BDI-II)^54–56^ and UPPS Impulsive Behaviour Scale (trait impulsivity).^57, 58^

Participants faced a computer screen, with both forearms resting on a table in a semi-flexed position. They performed a virtual 3D “Rolling Ball” game (Fig.1.A) previously used in several studies,^59–64^ including in PD patients;^38^ the response device was designed to detect horizontal index finger movements (Fig.1.B).^59, 63, 65^ Participants had to choose between a left or right index abduction, according to the position of a ball (preparatory cue) indicating the side for the required response; subjects needed to prepare their response in order to move as quickly as possible once a bridge (imperative signal) appeared. The arrival time (AT) corresponded to the time between the imperative and the moment the responding finger touched a metal contact. Responses of patients were classified according to PD dominance, i.e. depending on whether they were made with the most-affected (MA) or least-affected (LA) hand; these conditions were matched with the nondominant (ND) and dominant (D) hands of HC subjects, respectively, given that the MA side corresponded to the ND hand in most patients (13 of 20; Table 1).^38^

**Figure 1.**
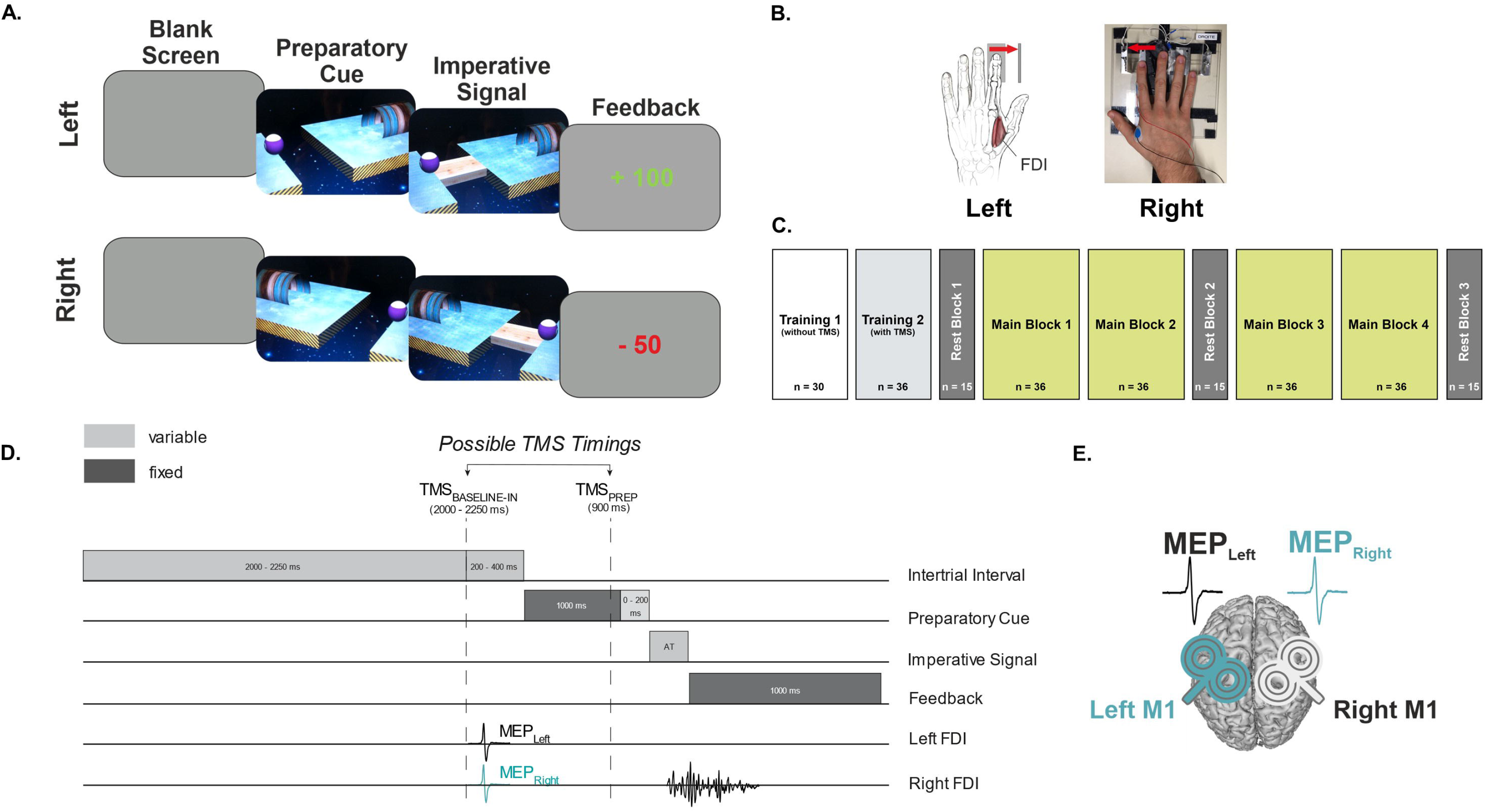
Experimental procedure. **A. Rolling ball task.** Participants performed an instructed-delay choice reaction time (RT) task, in which they had to choose between a left or right index finger abduction (left and right example shown) according to the position of a preparatory cue (a ball), separated from a goal by a gap. The side of the ball indicated the side of the required upcoming movement. Importantly, participants had to withhold their response until the onset of an imperative signal (a bridge), closing the gap. Once the bridge appeared, they had to respond as quickly as possible, with the correct finger, to make the ball roll over the bridge into the goal. Responding before the imperative signal caused the ball to fall into the gap. The imperative screen disappeared once a response was detected (or after a maximum time given to respond, individualised for each participant based on training performance) and a feedback was presented, in green or red, following a correct or incorrect answer, respectively. Following a correct response, participants received a positive score inversely proportional to the arrival time (AT) (the faster, the higher the score), ranging from 1 to 100 and depicted in green at the centre of the screen. Negative scores occurred when subjects responded too early (before the imperative signal), too late (more than the individualized maximum time) or with the incorrect finger. To prevent subjects from anticipating, we included catch trials (6 % of trials) in which the ball was not followed by a bridge; participants were instructed not to respond on these trials and were penalized if they still did. **B. Response device and EMG recording.** Each hand was positioned on a response device (left = graphic, right = photographic representation). Each device consisted of a pair of metal edges fixed on a plastic support, detecting the abduction movements of index fingers from the outer to the inner metal edge. Participants always needed to go back to the outer reference position before providing the next response. EMG activity was recorded from surface electrodes placed on the first dorsal interosseous (FDI) in both hands. **C. Experimental design.** After two training blocks, subjects performed four main blocks (*n* = 36 trials each), during which motor-evoked potentials (MEPs) were randomly elicited either at TMS_BASELINE-IN_ or TMS_PREP_. MEPs were also elicited in three rest blocks, outside the task context (TMS_BASELINE-OUT_). **D. Time course of a trial.** Each trial began with the preparatory cue displayed on the screen for 1000–1200 ms (purposely variable in order to decrease participants’ tendency to respond prematurely), followed by the imperative signal, remaining visible until a finger response was provided (within the maximum AT given to respond). Then, the feedback score based on the trial performance appeared for 1000 ms, followed by a blank screen (intertrial interval) lasting for 2200–2650 ms. TMS pulses were applied either during the intertrial interval (2000-2250 ms after the onset of the blank screen; TMS_BASELINE-IN_) or during the delay period (900 ms after the onset of the preparatory cue; TMS_PREP_). **E. TMS protocol.** Two small figure-of-eight coils were placed over both primary motor cortices for each participant, eliciting MEPs in the left and right hands with a 1 ms inter-pulse interval. (Figure adapted from Wilhelm et al., 2022)

Every participant performed the task in two sessions, with an identical block sequence (Fig.1.C). A first training block served to familiarise participants with the task (Training 1, no TMS), a second to calculate the individual median AT (Training 2, with TMS) (the latter was used to adapt the maximal time allowed to respond after the imperative signal (AT + 2 standard deviation [SD]) and to calculate feedback scores on correct trials).^64^ Next, the main phase of the experiment included four blocks of 36 trials, each consisting of an equal number of left- and right-hand trials, with TMS applied in 30/36 trials. TMS was delivered over both M1 at one of two possible timings (Fig.1.D) during the task: in some trials, to establish baseline measures of CSE, TMS occurred during the inter-trial interval (TMS_BASELINE-IN_; 10 trials/block), generating MEPs when participants were at rest within the task context. In other trials, TMS was applied between the preparatory cue and the imperative signal (TMS_PREP_; 20 trials/block), when patients were preparing a response either with their MA (RESP_MA_, 10 trials/block) or LA (RESP_LA_, 10 trials/block) hand (corresponding to RESP_ND_ or RESP_D_, respectively, in HC subjects). The remaining task trials (6/36) did not include TMS and served to prevent participants from anticipating a pulse at TMS_PREP_ whenever there had been none at TMS_BASELINE-IN_. Finally, we also acquired baseline MEPs at complete rest, outside the task, in front of a blank screen (TMS_BASELINE-OUT_; 3×15 trials/session) (Fig.1.C).

Electromyography (EMG) was used to measure the peak-to-peak amplitude of MEPs elicited in the First Dorsal Interosseous (FDI) muscle (task agonist), in the MA (MEP_MA_) and LA (MEP_LA_) hands of patients and the corresponding conditions in HC subjects (MEP_ND_ and MEP_D_, respectively). MEPs were also sorted according to the experimental condition in which they had been obtained (TMS_BASELINE-OUT_, TMS_BASELINE-IN_, TMS_PREP_ with RESP_MA/ND_ or RESP_LA/D_). Using EMG, we also extracted the RT in the task; the movement time (MT) was obtained by subtracting the RT from the AT in the corresponding trial. Details regarding the TMS protocol, EMG recording and data cleaning are presented in the Supplementary Material.

Differences in age and BDI-II between PD patients and HC subjects were evaluated using Mann-Whitney U-tests. Trait impulsivity was analysed with a multivariate analysis of variance (MANOVA) on scores reported at the four subscales of the UPPS questionnaire. In patients, total scores at the MDS-UPDRS part III as well as bradykinesia subscores were compared between the OFF- and ON-DBS sessions using a Wilcoxon test for each. We used Spearman’s rank correlations to check how disease duration (i.e. years since diagnosis) and time since surgery related to motor impairment based on total motor scores OFF-DBS.

Preparatory suppression was assessed by expressing MEPs acquired at TMS_PREP_ as percentage of MEPs elicited at TMS_BASELINE-IN_ in the corresponding hand. Group comparisons were then conducted using a four-way mixed repeated-measures ANOVA (RM-ANOVA) with GROUP (PD, HC) as between-subject factor and with DBS_CONDITION_ (ON, OFF), RESP_SIDE_ (MA/ND, LA/D) and MEP_SIDE_ (MA/ND, LA/D) as within-subject factors. Data was checked for sphericity using Mauchly tests whenever we conducted a RM-ANOVA. Moreover, to assess the degree of preparatory suppression in each subcondition, paired *t*-tests were carried out to compare the raw MEP values (mV) at TMS_PREP_ to those at TMS_BASELINE-IN_. Next, we used Spearman’s rank correlations to check for a possible relationship between levels of preparatory suppression and disease duration, time since surgery as well as bradykinesia scores (in the same hand) in PD patients. For raw measures of CSE and behaviour during the task, see Supplementary Material. Analyses were carried out using JASP (version 0.16.4.0; JASP Team, 2022). For all RM-ANOVAs, we ran post-hoc comparisons using Tukey’s HSD. Data is shown as mean ± SD; statistical significance was set at P < 0.05. For multiple comparisons, we used FDR corrections.

Data supporting the present study findings are available upon request to the authors.

## Results

A summary of group comparisons is shown in Table 2. PD patients and HCs did not differ in age, but patients had higher depression scores at the BDI-II, as expected. Both groups also displayed similar global scores of trait impulsivity at the UPPS questionnaire (λ_4,35_ = 0.79; *P* = 0.07), although PD patients exhibited less perseverance (*P* < 0.01) and tended to show less premeditation (*P* = 0.08). Patients scored significantly higher on the MDS-UPDRS-III (total and bradykinesia), OFF-DBS compared to ON-DBS (both *P* < 0.001), confirming the alleviation of motor symptoms in the presence of STN stimulation. Contrary to our past study, in this advanced cohort, total clinical scores did not correlate positively with disease duration OFF-DBS (*P* = 0.1); yet, they did so with time since surgery (*rho* = 0.47, uncorrected *P* = 0.035, corrected *P* = 0.12).

**Table 2.**
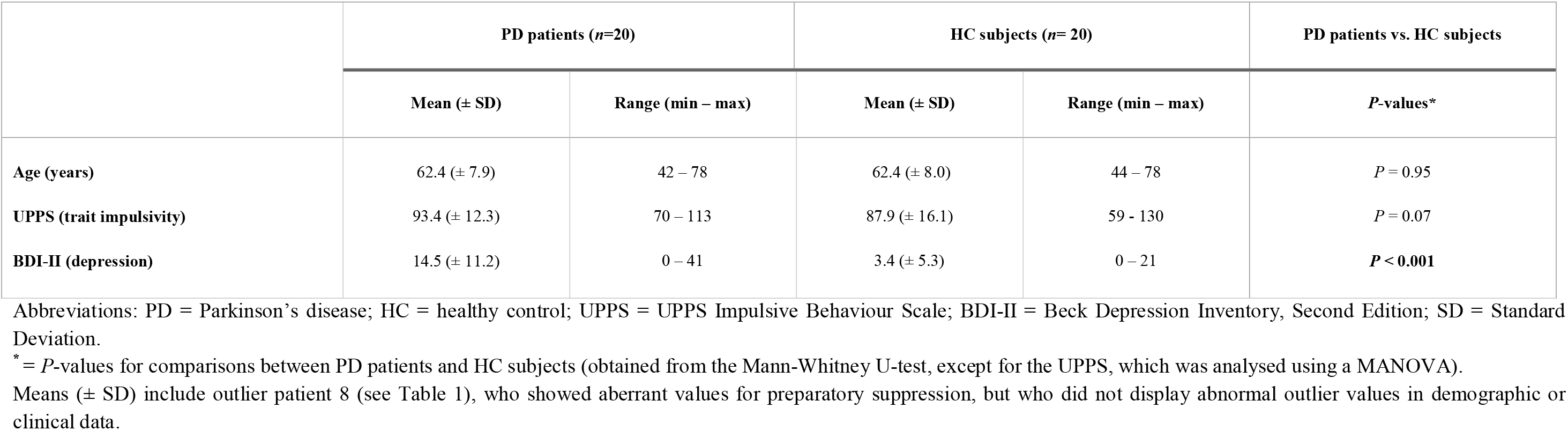
Demographic and clinical data in PD patients and HC subjects.

|  | PD patients (*n*=20) |  | HC subjects (*n*= 20) |  | PD patients vs. HC subjects |
| --- | --- | --- | --- | --- | --- |
| | Mean (± SD) | Range (min – max) | Mean (± SD) | Range (min – max) | *P*-values* |
| Age (years) | 62.4 (± 7.9) | 42 – 78 | 62.4 (± 8.0) | 44 – 78 | *P* = 0.95 |
| UPPS (trait impulsivity) | 93.4 (± 12.3) | 70 – 113 | 87.9 (± 16.1) | 59 - 130 | *P* = 0.07 |
| BDI-II (depression) | 14.5 (± 11.2) | 0 – 41 | 3.4 (± 5.3) | 0 – 21 | *P* < **0.001** |

Abbreviations: PD = Parkinson's disease; HC = healthy control; UPPS = UPPS Impulsive Behaviour Scale; BDI-II = Beck Depression Inventory, Second Edition; SD = Standard Deviation.
\* = *P*-values for comparisons between PD patients and HC subjects (obtained from the Mann-Whitney U-test, except for the UPPS, which was analysed using a MANOVA).

The analysis of preparatory suppression revealed one patient with aberrant values (patient 8 in Table 1), who was identified as an extreme outlier, based on the IQR rule (inset Fig.2);^66, 67^ this patient and the corresponding control subject were subsequently removed from MEP analyses.

**Figure 2.**
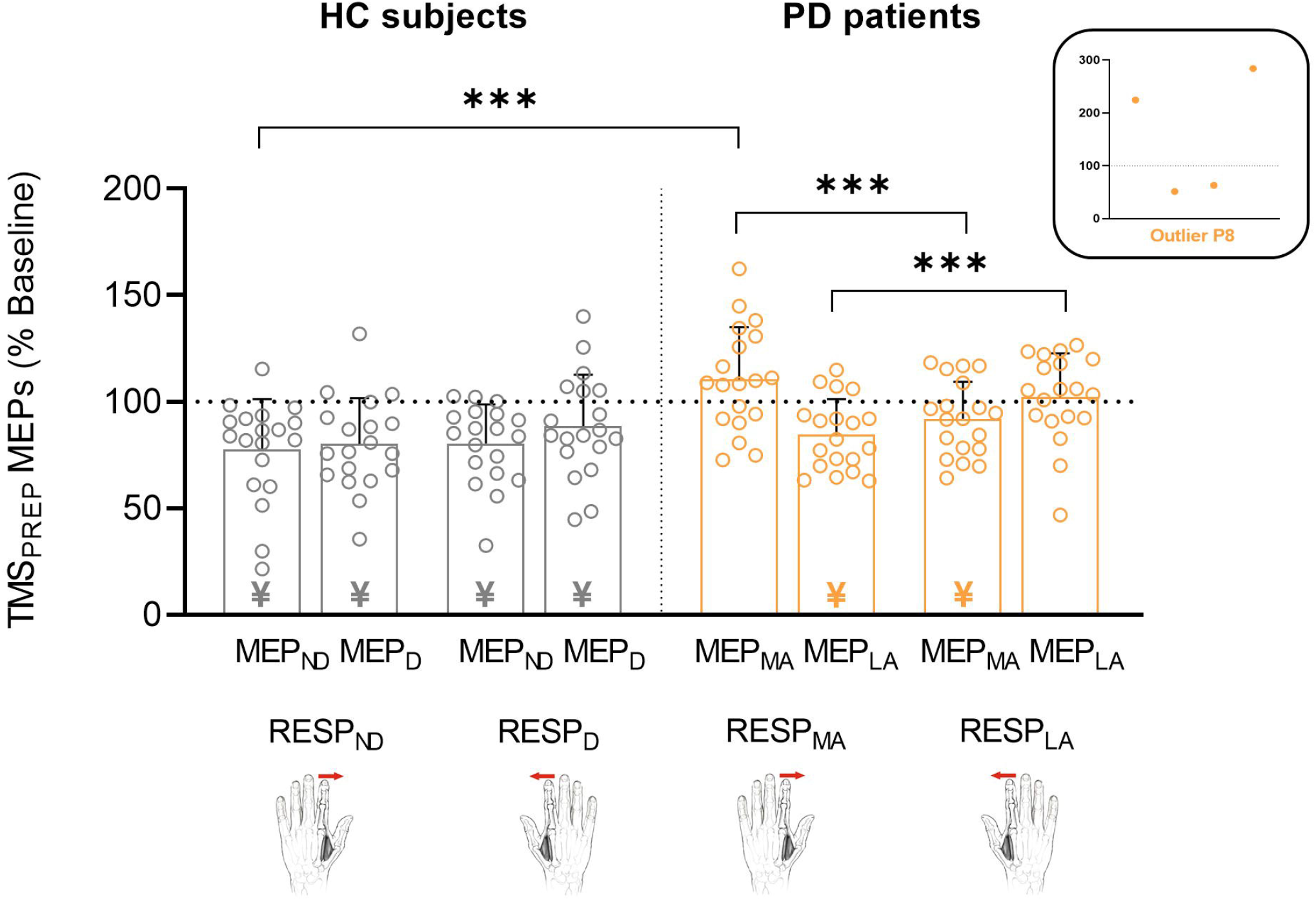
Preparatory suppression in Parkinson’s disease (PD) patients (n=19; in orange) and healthy control (HC) subjects (n=19; in grey), after exclusion of outlier patient P8 (see Table 1) and the corresponding control subject. Note that for every participant, results of both DBS sessions are pooled together. Each individual dot represents data for a single subject, while the bar plot represents the mean value (+ SD) for the condition. Levels of preparatory suppression were obtained by expressing amplitudes of motor-evoked potentials (MEPs) measured at TMS_PREP_ as a percentage of those elicited at TMS_BASELINE-IN_, shown for the most-affected (MEP_MA_) and least-affected (MEP_LA_) hands in PD patients, which corresponded to the non-dominant (MEP_ND_) and dominant (MEP_D_) sides in control subjects. The hand figures represent the responding side (RESP_SIDE_); for the purpose of illustration, in patients, the left and right hands represent MA and LA hand responses (RESP_MA_ and RESP_LA_), respectively. Thus, for each group, the bars represent MEPs according to the hand in which they were measured and to the hand that was responding (two peripheral bars) or not (two central bars) in the trial. The inset shows the values of outlier patient P8 (the inset’s axes represent the same variables and conditions as the barplot). Taken together, this figure shows consistent preparatory suppression in all conditions in HC subjects, but unveils a lack of this mechanism in the responding hand conditions in PD patients, particularly on the MA side, which significantly differed from the corresponding condition in HC subjects (*P* < 0.001; all other *P* > 0.49). Paired t-tests: ¥ = MEPs probed at TMS_PREP_ significantly different from those probed at TMS_BASELINE-IN_; mixed RM-ANOVA: *\*\*\*P* < 0.001.

The RM-ANOVA run on the remaining percentage MEP data (n=19 per group) revealed a significant effect of the factor GROUP [*F*(1,36) = 9.2, *P* < 0.01] due to larger values in PD patients (97.4 ± 27.4 %) compared to HC subjects (81.7 ± 24.6 %), indicating overall less preparatory suppression in the former, consistent with our recent findings on a separate sample of earlier-stage PD patients.^38^

Also in line with this past study, analyses yielded a significant GROUP x RESP_SIDE_ x MEP_SIDE_ interaction [*F*(1,36) = 13.6, *P* < 0.001]: as shown in Figure 2, healthy participants exhibited similar levels of MEP suppression during action preparation no matter the hand considered for MEPs (MEP_ND_ or MEP_D_) or the hand required for the upcoming movement response (RESP_ND_ or RESP_D_; all *P* > 0.42 between conditions). Accordingly, FDR-corrected paired *t*-tests run between raw MEP values (mV) at TMS_PREP_ and those at TMS_BASELINE-IN_ were all significant (all *t*(18) > 3.44, all *P* < 0.01), confirming the systematic presence of preparatory suppression in HC subjects, as repetitively shown in past studies.^9, 30, 33, 38, 62^ In contrast, in PD patients, post-hoc analyses showed that preparatory suppression depended on the condition: corticospinal suppression was weaker when MEP_MA_ and MEP_LA_ were measured in the hand selected for the forthcoming movement (i.e. RESP_MA_ and RESP_LA_ trials, respectively) compared to when these MEP_MA_ and MEP_LA_ occurred on the non-responding hand side (i.e. in RESP_LA_ and RESP_MA_ trials, respectively; both *P* < 0.001). FDR-corrected paired *t*-tests between raw MEPs at TMS_PREP_ and TMS_BASELINE-IN_ further supported a lack of preparatory suppression on the responding side (both *t*(18) < −3.03, both *P* > 0.16), with percentage values even leaning visually towards facilitation, particularly in the MA hand; on the contrary, MEPs were still suppressed on the non-responding side (both *t*(18) > 2.59, both *P* < 0.02). Consistent with these findings, when comparing levels of suppression between groups, a significant difference emerged between the MEP_MA_–RESP_MA_ condition in patients and the corresponding trials in HC subjects (i.e., MEP_ND_–RESP_ND_, Fig.2). Importantly, the equivalent group comparison for the MEP_LA_–RESP_LA_ condition was not significant (*P* = 0.49), suggesting a more pronounced deficit of preparatory suppression on the PD-dominant side, similar to our past study on earlier-stage patients.^38^

The RM-ANOVA showed no overall effect of DBS on corticospinal suppression in patients, as illustrated by the absence of a GROUP x DBS interaction (*F*(1, 36) = 3.23, *P* = 0.08). Nonetheless, it revealed a significant GROUP x DBS x MEP_SIDE_ interaction (*F*(1, 36) = 5.08, *P* = 0.03), for which post-hoc analyses, however, unveiled no significant effect of the stimulation (Supplementary Fig.1). All other interactions involving the factor DBS were non-significant (all *P* > 0.81). Bayesian equivalents of the RM-ANOVA^68^ for measures of preparatory suppression were in line with an absence of effect of STN-DBS (see Supplementary Material).

Finally, in PD patients, we ran Spearman’s rank correlations between clinical results (OFF-DBS) and preparatory suppression (in the RESP_MA_-MEP_MA_ condition, given its most pronounced deficit); for the latter, results from ON- and OFF-DBS sessions were pooled, considering the absence of effect of stimulation. Given that time since surgery appeared to reflect global motor progression, all correlations that did not involve this variable directly, were corrected for it. Neither clinical time variables nor bradykinesia scores correlated with preparatory suppression in the RESP_MA_-MEP_MA_ condition (all corrected *P* > 1.0). Given the existence of positive correlations in our previous PD cohort, we therefore extended the analyses to the LA hand; interestingly, here, preparatory suppression in the RESP_LA_-MEP_LA_ condition correlated with bradykinesia in the same hand (*rho* = 0.56, uncorrected *P* = 0.017, corrected *P* = 0.085; Fig.3.A). Given this significant result limited to the LA hand, we also verified how preparatory suppression in the RESP_MA_-MEP_LA_ (i.e. non-responding) condition evolved with time variables; this revealed a significant correlation with time since surgery (*rho* = −0.58, uncorrected *P* = 0.009, corrected *P* = 0.09; Fig.3.B).

**Figure 3.**
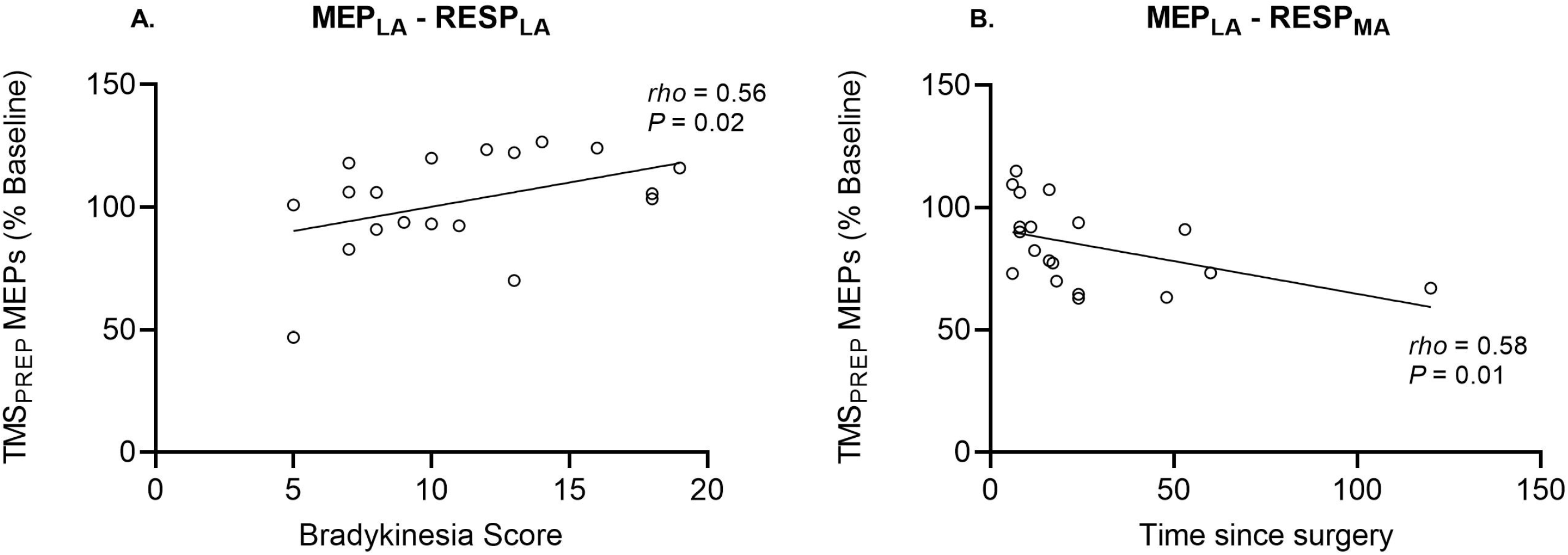
Spearman’s rank correlations in PD patients. shown for preparatory suppression (i.e. percentage MEPs at TMS_PREP_) on the least-affected (**MEP_LA_**) side when the latter was **A.** responding in the task (**RESP_LA_**) in relation to **bradykinesia scores** in the LA hand (based on the MDS-UPDRS-III) *off-*DBS (and corrected for time since surgery) or **B.** not responding in the task (**RESP_MA_**) in relation to **time since surgery** (in months). Note that outlier patient P8 was excluded from these correlations. In the absence of an effect of DBS, data from both sessions are pooled together. Note the positive relationship between MEP_LA_-RESP_LA_ preparatory suppression and bradykinesia scores, when correcting for time since surgery (*rho* = 0.56, uncorrected *P* = 0.017, FDR-corrected *P* = 0.085), and the negative relationship between MEP_LA_-RESP_MA_ preparatory suppression and time since surgery (*rho* = −0.58, uncorrected *P* = 0.009, FDR-corrected *P* = 0.09).

## Discussion

In order to investigate a possible involvement of the STN in shaping corticospinal dynamics during movement preparation in advanced PD, we applied TMS over M1 during an instructed-delay choice RT task in patients implanted with bilateral STN-DBS. Contrary to HCs, those patients exhibited a lack of corticospinal suppression during action preparation, a deficit limited to the responding hand and most predominant on the MA side, consistent with prior findings in earlier-stage patients. However, contrary to the latter, here significant correlations between preparatory suppression and clinical variables were found only for the LA hand. Importantly, deficits of preparatory suppression were not restored ON-DBS.

Our findings in healthy subjects are in line with several past studies reporting a strong suppression of CSE during movement preparation, no matter if MEPs are acquired in the responding or non-responding hand and no matter if the D or ND side is probed.^9, 30, 31, 38, 64, 65, 69^ Patients with advanced PD, however, displayed an alteration of this mechanism, as MEPs consistently lacked suppression in the responding hand; CSE even appeared facilitated when considering responses on the MA side. Of importance is that specifically in the latter condition, preparatory suppression differed from control subjects, in line with our findings in non-operated PD patients.^38^

Surprisingly, we did not observe any difference in the degree to which MEPs were suppressed between the OFF- and ON-DBS sessions, meaning preparatory suppression deficits were present even with the stimulator ON. Since we found highly significant improvements in UPDRS scores ON-DBS – consistent with previous literature^17, 27, 70, 71^ – it is unlikely that the absence of difference in preparatory suppression was caused by residual effects of DBS or by excessive drug-induced stimulation of the dopaminergic system with the stimulator OFF.^18, 19, 38^ Even if the absence of effect of STN-DBS on corticospinal suppression can appear surprising, given the *a priori* plausible role of the STN in such phenomenon,^8, 72^ this result is in line with the previously demonstrated lack of modulation of preparatory suppression by DRT.^38^ Thus, despite the involvement of BG in preparing externally-triggered movements^17, 73^ and of dopamine in gain modulation,^74–77^ this suggests that neural pathways underlying corticospinal suppression during action preparation could be non-dopaminergic and lay outside the motor-STN or even BG circuits. Other neurotransmitter systems might be involved in this CSE regulation, with the noradrenergic^78^ one as a potential candidate. Its role has been amply described in cognition,^78^ and the activity of the locus coeruleus (LC) – the principal source of noradrenaline – has been shown to regulate the signal-to-noise ratio both at the level of the LC and target neurons.^79^ Interestingly, in PD, LC pathology can occur before losses in other neurotransmitter systems.^80^ Therefore, the LC might be involved in modulating levels of preparatory suppression, which should warrant future investigations in that direction. Note, however, that dopaminergic and noradrenergic systems could both be involved in such corticospinal suppression, possibly explaining why DRT or STN modulation alone did not restore deficits in PD patients. Another plausible explanation for our findings is that the lack of preparatory suppression in PD actually reflects adaptive compensatory mechanisms along the motor network, in order to increase M1 excitability,^81–83^ as previously suggested.^38^

In this STN-DBS cohort, contrary to our previous findings in earlier-stage PD,^38^ we did not find significant correlations between levels of preparatory suppression and disease duration; this appears in line with the concomitant absence of correlation between disease duration and motor impairment. The latter could be due to the surgical intervention itself, acting as disruptor in the natural disease progression.^84–87^ Interestingly, in the present study, the mean age at diagnosis was a decade below the one of our previous cohort, and past studies have shown quicker motor progression with higher age at disease onset.^88, 89^ The presence of DRT in both sessions could furthermore have influenced the relationship between disease duration and motor impairment, as the OFF-medication state is typically a better indicator of motor symptom progression.^88, 90–92^ Time since surgery, however, did correlate with total UPDRS scores OFF-DBS, indicating a progression of the disease after the surgical intervention, consistent with literature.^93, 94^ Moreover, we did find a link between preparatory suppression and bradykinesia scores in the LA hand (when accounting for time since surgery), contrary to previous results uncovering such a link for the MA hand.^38^ Interestingly, based on the clinical asymmetry characterising PD, previous studies already suggested that the latter evolves at two different rates of progression in both cerebral hemispheres: those are considered to represent two different disease stages,^95–98^ with the hemisphere of the clinically LA side exhibiting an “earlier” stage. Compensatory processes in neurodegenerative disorders – although occurring on many different temporo-spatial scales – tend to decline with disease progression and may be replaced by maladaptive reorganization; thus, compensatory changes are often argued to be less likely in more advanced PD.^81, 99^ Therefore, it is plausible that the LA hemisphere still shows a capacity for adaptive changes, as also suggested by the presence of a significant correlation between time since surgery and MEP suppression in the LA, non-responding hand. In contrast, measures on the MA side (considered to represent a “later” stage), may indicate the presence of maladaptive changes, after a loss of compensatory faculties.

Taken together, two non-exclusive hypotheses emerge to explain why, overall, treatment – whether DRT or STN-DBS – did not restore preparatory suppression in our studies. On the one hand, it could suggest the involvement of structures other than BG/STN (e.g. LC), possibly linked to alterations of non-dopaminergic systems (e.g. noradrenergic). On the other hand, compensatory mechanisms could be at play, with the MA and LA hemispheres appearing to be at two different disease stages and thus displaying more or less maladaptive changes, respectively. Nevertheless, as true for many progressive disorders, the study of PD is characterized by the difficulty to disentangle direct disease effects and compensatory/maladaptive changes developing over time.^82^ Therefore, the longitudinal nature inherent to both of these processes renders interpretations based on cross-sectional data more complicated, particularly in advanced PD cohorts treated with STN-DBS, who are likely characterised by both of these influences.^100^

In conclusion, our study provides first insights into alterations of corticospinal dynamics underlying movement preparation in advanced PD patients treated with bilateral STN-DBS, by uncovering a clear deficit in corticospinal suppression typically shaping neural activity during voluntary action preparation in healthy subjects. Such deficits were limited to the responding hand side and most pronounced in the MA one, consistent with previous findings in earlier-stage patients treated with DRT only. Contrary to the latter cohort, however, preparatory suppression appeared to be related to clinical variables for the LA hand only, hinting at a sequential disease-stage process – possibly linked to a progressive shift from compensatory to maladaptive changes – in the LA and MA hemispheres, respectively. Even though STN-DBS improved motor impairment, it did not restore preparatory suppression in this cohort, similar to the absence of effect of DRT in our recent study in earlier-stage PD. As such, the question regarding which structure(s) may be involved in generating preparatory suppression and the cause of its consistent disruption in PD patients, remains open. Future studies should consider the possibility that structures beyond the STN and non-dopaminergic neurotransmitter systems may contribute to shape corticospinal dynamics during action preparation.

## Data Availability

Data supporting the present study findings are available upon reasonable request to the authors.

## Abbreviations

CSE: corticospinal excitability
DBS: deep brain stimulation
DRT: dopamine replacement therapy
FDI: first dorsal interosseous
HC: healthy control
M1: primary motor cortex
MDS-UPDRS-III: Movement Disorder Society Unified Parkinson’s Disease Rating Scale, Part III
MEP: motor evoked potential
PD: Parkinson’s disease
STN: subthalamic nucleus
TMS: transcranial magnetic stimulation

## Acknowledgements

First, we are very grateful to all patients and control subjects who participated in this research project, which – for many of them – took place during a complicated sanitary context. We would also like to thank Prof. Ivanoiu, Dr. Leempoel, Dr. Peeters, Dr. Verougstraete (Saint-Luc University Hospital, Brussels, Belgium) and Dr. Boogers (UZ Leuven, Leuven, Belgium) for their valuable help in patient recruitment, as well as Marie-Lou Hoang and Zélie Rosselli for their relevant contribution to participant recruitment and data collection. Next, we are thankful to the Statistical Methodology and Computing Service of the *UCLouvain*, as well as to Dr. Foffani (CINAC, *Hospital Universitario HM Puerta del Sur*, Madrid, Spain) for his valuable comments on an earlier version of the manuscript. This paper is dedicated to Prof. Jeanjean, former head of the Adult Neurology Department and renowned movement disorder specialist at the Saint-Luc University Hospital, as well as former clinical mentor of author EW. She passed away on 15th January 2020.

## Supplementary material

Supplementary material is available online.

## Author contributions

Conceptualisation and Methodology: E.W., G.D., C.Q. and J.D.; Participant Recruitment: E.W. and I.C.; Investigation and Data Curation: E.W. and I.C.; Formal Analysis: E.W. and S.P.; Data Interpretation: E.W., G.D., C.Q., S.P. and J.D; Writing - Original Draft: E.W.; Writing – Review & Editing: E.W., G.D., C.Q., I.C., S.P., C.R., B.N. and J.D.; Visualisation: E.W.; Funding Acquisition: E.W., G.D., C.Q. and J.D.; Supervision: J.D.

