## Supplementary Material for "Subthalamic deep brain stimulation alleviates motor symptoms without restoring deficits in corticospinal suppression during movement preparation in Parkinson’s disease"

### Supplementary methods

**Experimental protocol**

#### **TMS and EMG recording**

Monophasic pulses were delivered using a double-coil TMS method whereby pulses are delivered over the two M1 with a 1 ms inter-pulse interval, as used in many previous studies, including in PD patients.^1-6^ Pulses were generated with two small figure-of-eight coils (wing internal diameter: 35 mm). The coil stimulating the right M1 was connected to a Magstim 200^2^ magnetic stimulator (Magstim, Whitland, Dyfed, UK) and the coil stimulating the left M1 to a Magstim BiStim^2^ magnetic stimulator. Both coils were placed tangentially over both M1 with the handle pointing backward and laterally at a 45° angle away from the midline (Fig.1.E, main manuscript), approximately perpendicular to the central sulcus, in order to induce a posterior-anterior current in the underlying neural cells.^7, 8^

For each M1, we identified the optimal scalp position for eliciting contralateral MEPs in the First Dorsal Interosseus (FDI; task agonist), as well as in the *Abductor Digiti Minimi* (ADM) and *Abductor Pollicis Brevis* (APB) muscles; this multi-muscle hotspot was marked on a cap placed on the participant’s head to provide a reference mark throughout the experiment.^9^ For each M1, the resting motor threshold was determined as the minimal TMS intensity required to evoke MEPs of minimum 50 μV peak-to-peak in each of the three muscles in at least 5 out of 10 consecutive trials.^10^ For each hemisphere, the intensity of TMS throughout the experiment was always set at 115% of the individual resting motor threshold,^11^ which allowed us to elicit reliable FDI, ADM and APB MEPs bilaterally. Notably, MEPs elicited in the ADM and APB were obtained as part of another study; here we focus on the FDI, the prime-mover in the task.

EMG activity was recorded from the FDI using surface electrodes (Ambu BlueSensor NF-50-K/12/EU, Neuroline, Medicotest, Oelstykke, Denmark) placed over the left- and right-hand muscles (Fig.1.B, main manuscript). EMG data was collected for 3000 ms on each trial, starting 200 ms before the TMS pulse; signals were monitored visually on a computer screen during the whole experiment. The raw EMG signals were amplified (gain of 1K), band-pass filtered on-line (10-500 Hz), notch-filtered (50 Hz) (Digitimer D360, Hertfordshire, UK) and digitized at a sampling rate of 2 kHz (CED 1401-3 ADC12 and Signal 6 software, Cambridge Electronic Design Ltd) for off-line analysis. Data extraction and cleaning were performed using fully automated procedures in Signal 6 and R Studio (https://www.r-project.org/), respectively.

Trials with any background EMG activity > 3 SD above the mean of the root mean square in the 200-ms window before the TMS pulse were removed from the analysis. Note that we also excluded trials in which subjects had made an error (e.g. responding with incorrect finger); these were rare given the task simplicity. Then, for each muscle and condition, we excluded trials with peak-to-peak MEP amplitudes larger or smaller than 3 SD around the mean. The EMG recording allowed to extract the RT in the task, defined as the moment after the imperative signal when the Hjorth activity of the EMG envelope was above 0.2.^12, 13^

#### **Tremor management in PD patients**

We carefully chose our patients to be non-dyskinetic and non-tremor dominant at the moment of the experiment. Nevertheless, when the patients arrived for their first day of testing, a few of them did present slight signs of tremor. When present, however, this tremor was mild (i.e. no patient had a MDS-UPDRS-III tremor subscore higher than 1) and was most frequently of postural nature. In some patients, a stress-related, intermittent rest tremor was visible upon installing them for the TMS experiment, before the initiation of the task (i.e. while searching for the hotspots and determining rMTs). By visually tracking the EMG window on the computer screen, we would then regularly, but gently remind them to relax as much as possible, while also making sure that their hand, arm and back positions were as comfortable and relaxed as possible, which worked well. Then, during the task execution itself (i.e. action), tremor was practically absent in all patients (typical of PD). Finally, we also cleaned all trials based on EMG background activity before the TMS pulse, in order to prevent any possible intermittent tremor-related activity from contaminating our EMG data (cleaning based on the root mean square in the 200 ms window preceding the TMS pulse, see above).

#### **Bradykinesia scores based on the MDS-UPDRS-III**

In each patient, we more specifically quantified bradykinesia, by selecting a subset of items more closely reflecting this cardinal motor symptom. Based on an inconsistent establishment of bradykinesia subscores in the literature,^14-16^ here, we included items 3.2, 3.4 - 3.8 and 3.14 of the MDS-UPDRS-III, in order to englobe the widest set of motor characteristics related to this cardinal symptom.^17^

**Statistical analysis**

**Raw measures of corticospinal excitability**

Next, we investigated whether the two groups displayed differences in resting motor thresholds by means of a three-way mixed RM-ANOVA with GROUP (PD, HC) as between-subject factor and with DBS_CONDITION_ (ON, OFF) and MEP_SIDE_ (MA/ND, LA/D) as within-subject factors. Then, we focused on measures of CSE at rest by considering MEPs acquired at TMS_BASELINE-OUT_ (i.e. outside the main blocks) and at TMS_BASELINE-IN_ (i.e. within the main blocks). The raw amplitude of these MEPs (mV) was analysed using a four-way mixed RM-ANOVA, with GROUP (PD, HC) as the between-subject factor and with DBS_CONDITION_ (ON, OFF), TMS_BASELINE_ (OUT, IN) and MEP_SIDE_ (MA/ND, LA/D) as within-subject factors.

**Behaviour during the rolling ball task**

In order to analyse the behaviour of participants during the task, we focused on trials in which TMS pulses were applied during the inter-trial interval (TMS_BASELINE-IN_) or trials in which there was no TMS pulse at all, in order to avoid a possible disturbance effect of the pulse;^2, 18, 19^ all those trials were pooled together. For the analyses of RT and MT, we conducted two separate three-way mixed RM-ANOVAs, with GROUP (PD, HC) as between-subject factor and with DBS_CONDITION_ (ON, OFF) and RESP_SIDE_ (MA/ND, LA/D) as within-subject factors.

**Bayesian analyses on measures of preparatory suppression**

We ran an additional Bayesian equivalent of the frequentist RM-ANOVA with JASP. In this case, the Bayes factor (BF10) quantifies the evidence for the alternative hypothesis against the null hypothesis and the prior and posterior inclusion probabilities [P(incl) and P(incl|data)] refer to the importance of each parameter based on the prior and posterior probabilities of each model including it, respectively.

Bayesian analyses validated the findings of the frequentist approach reported in the main manuscript, by showing very strong evidence for a GROUP effect (BF10 = 54.9) as well as a GROUP x RESP_SIDE_ x MEP_SIDE_ interaction (BF10 = 55.2).

| **Analysis of Effects** | | | | | | | | | | |
| --- | --- | --- | --- | --- | --- | --- | --- | --- | --- | --- |
| **Effects** | | **P(incl)** | | **P(excl)** | | **P(incl\|data)** | | **P(excl\|data)** | | **BF_incl_** |
| GROUP |  | 0.886 |  | 0.114 |  | 0.998 |  | 0.002 |  | 54.932 |
| DBS x GROUP |  | 0.503 |  | 0.497 |  | 0.363 |  | 0.637 |  | 0.563 |
| DBS x RESP_SIDE_ x GROUP |  | 0.120 |  | 0.880 |  | 0.017 |  | 0.983 |  | 0.130 |
| DBS x MEP_SIDE_ x GROUP |  | 0.120 |  | 0.880 |  | 0.108 |  | 0.892 |  | 0.890 |
| RESP_SIDE_ x   MEP_SIDE_ x GROUP |  | 0.120 |  | 0.880 |  | 0.883 |  | 0.117 |  | 55.241 |
| DBS x RESP_SIDE_ x MEP_SIDE_ x GROUP |  | 0.006 |  | 0.994 |  | 3.169×10^-4^ |  | 1.000 |  | 0.053 |

As for a possible effect of the DBS_CONDITION_, Bayesian analyses showed:

- DBS x GROUP: anecdotal evidence in favour of the null hypothesis
- DBS x RESP_SIDE_ x GROUP: moderate evidence in favour of the null hypothesis
- DBS x MEP_SIDE_ x GROUP: anecdotal evidence in favour of the null hypothesis
- DBS x RESP_SIDE_ x MEP_SIDE_ x GROUP: strong evidence in favour of the null hypothesis

Overall, this shows a tendency towards evidence for the null hypothesis, with interactions involving the DBS_CONDITION_ revealing Bayes factors BF10 of up to 0.05. These results are in line, again, with the findings reported in the main manuscript, which are in favour of an absence of effect of STN stimulation on measures of corticospinal suppression in the current cohort of advanced PD patients treated with bilateral STN-DBS.

**Supplementary results**

**Raw measures of corticospinal excitability**

The three-way RM-ANOVA performed on resting motor thresholds did not reveal any significant effect of the main factor GROUP [*F*(1,36) = 0.94, *P* = 0.34] (mean: 46 ± 8% and 48 ± 9%, in PD and control subjects, respectively) nor did it show a significant GROUP x DBS interaction [*F*(1,36) = 0.04, *P* = 0.84], indicating the absence of an effect of the stimulation on resting motor thresholds in patients, consistent with previous studies.^20, 21^ Furthermore, we did not find any significant effect of the factor MEP_SIDE_ [*F*(1,36) = 3.59, *P* = 0.07] nor a significant GROUP x MEP_SIDE_ interaction [F(1,36) = 1.43, *P* = 0.24].

When considering MEPs acquired at rest (TMS_BASELINE-OUT_ and TMS_BASELINE-IN_, i.e. outside and within task blocks), always at 115% of resting motor threshold, the four-way RM-ANOVA showed a significant effect of the factor BASELINE [*F*(1,36) = 63.03, *P* < 0.001], without yielding a significant effect of the factor GROUP [*F*(1,36) = 0.38, *P* = 0.54] nor of the GROUP x BASELINE interaction [*F*(1,36) = 0.76, *P* = 0.39]. Post-hoc analyses revealed that both groups exhibited larger MEP amplitudes at TMS_BASELINE-IN_ (2.36 mV ±1.7 and 1.97 mV ±1.4, for patients and controls respectively) relative to TMS_BASELINE-OUT_ (1.29 mV ±1.1 and 1.17 mV ±1.0, for patients and controls respectively). This reflects an increase of CSE within the task context, which is in line with previous findings in healthy subjects and PD patients.^2, 3^ The RM-ANOVA also yielded a significant GROUP x BASELINE x MEP_SIDE_ interaction [*F*(1,36) = 4.24, *P* = 0.047]. Post-hoc analyses revealed that – independent of the DBS condition – for TMS_BASELINE-IN_, PD patients exhibited significantly larger MEPs in the MA hand (MEP_MA_) compared to the LA hand (MEP_LA_) (P < 0.05), whereas HC subjects rather displayed larger MEPs in the D hand (MEP_D_) compared to the ND hand (MEP_ND_) (P < 0.01); such differences did not exist at TMS_BASELINE-OUT_ (both P > 0.13). Finally, we did not detect any effect of DBS on rest MEPs in PD patients, as illustrated by the absence of a significant GROUP x DBS interaction [*F*(1,36) = 0.32, *P* = 0.58] or of any other interaction involving the factor DBS (all *P* > 0.08).

The fact that raw MEP measures at TMS_BASELINE-IN_ in our PD cohort did not vary according to the DBS status, despite significantly higher CSE in the MA compared to the LA hand, reinforces the idea of possible compensatory mechanisms. Indeed, previous studies already showed an absence of normalisation of increased MEPs through STN stimulation in advanced PD,^20, 22^ suggesting that compensatory influences may not be simply replaced by the effects of STN-DBS, even if motor symptoms are well improved.^23^

**Behaviour during the rolling ball task**

The three-way RM-ANOVAs revealed a significant effect of the main factor GROUP for RT [*F*(1,36) = 21.09, *P* < 0.001], but not MT [*F*(1,36) = 0.54, *P* = 0.47] data, hence illustrating overall slower movement initiation but not execution in PD patients compared to HC subjects in the present cohort (mean RT: 344.2 ± 130.2 ms and 231.8 ± 57.4 ms, in PD and HC subjects respectively; mean MT: 91.4 ± 66.1 ms and 87.5 ± 24.5 ms, respectively) (Supplementary Fig. 2). Such results did not depend on the disease or hand dominance, as revealed by the absence of a GROUP x RESP_SIDE_ effect on RT and MT values [both *F*(1,36) < 0.28, both *P* > 0.60], neither did they depend on the DBS status [both *F*(1,36) < 0.24, both *P* > 0.3].

Despite longer RTs in the patient group, in line with increased cognitive slowing with disease progression,^24, 25^ we observed no effect of DBS on patients’ task behaviour, similar to neurophysiological results, but contrary to clinical motor scores. In other studies, STN-DBS has been shown to increase movement velocity,^26-28^ and shorten RTs,^28-30^ but the latter finding has been demonstrated to depend on task complexity;^31^ it should be noted that none of those tasks are directly comparable to the one used in the current project. The absence of an effect of group and of STN-DBS on movement times in our task, in particular, could potentially be linked to a limited abduction amplitude missing slight variations in movement vigour; indeed, our task was developed specifically to allow investigating preparatory suppression, and hence wasn’t purposefully designed to serve as a tool to acquire kinematic measures of bradykinesia.

**Supplementary Table. Average number of trials remaining per condition after data cleaning in PD patients and HC subjects.**

|  | PD patients | | | | HC subjects | | | |
| --- | --- | --- | --- | --- | --- | --- | --- | --- |
|  | **MEP_MA_** | | **MEP_LA_** | | **MEP_ND_** | | **MEP_D_** | |
| **TMS_BASELINE-OUT_** | 41.6 (± 4.6) | | 41.3 (± 4.5) | | 42.2 (± 3.3) | | 41.6 (± 3.6) | |
| **TMS_BASELINE-IN_** | 25.7 (± 6.2) | | 25.8 (± 6.2) | | 27.4 (± 4.9) | | 27.5 (± 4.7) | |
| **TMS_PREP_** | RESP_MA_ | RESP_LA_ | RESP_MA_ | RESP_LA_ | RESP_ND_ | RESP_D_ | RESP_ND_ | RESP_D_ |
|  | 24.8 (± 6.6) | 25.5 (± 7.0) | 24.9 (± 6.8) | 25.6 (± 7.1) | 29.0 (± 5.7) | 29.6 (± 5.7) | 29.0 (± 5.6) | 29.6 (± 5.7) |

PD = Parkinson’s disease; TMS_BASELINE-OUT_ = pulse delivered outside the context of the task, at complete rest in front of a blank screen; TMS_BASELINE-IN_ = pulse delivered within the context of the task, during the inter-trial interval; TMS_PREP_ = pulse delivered during the preparatory phase, between the appearance of the preparatory cue and the imperative signal; MEP_MA_ and MEP_LA_ = motor evoked potentials probed in the most- and least-affected hand, respectively; RESP_MA_ and RESP_LA_ = most- and least-affected hand selected for the upcoming movement response, respectively. Note that the MA and LA conditions in PD patients were considered comparable to the non-dominant (MEP_ND_, RESP_ND_) and dominant (MEP_D_, RESP_D_) conditions in HC subjects, respectively. Means (± SD) include outlier patient P8, who showed aberrant values for preparatory suppression, but who did not display abnormal outlier values in demographic, clinical or behavioural task data.

### Supplementary figures

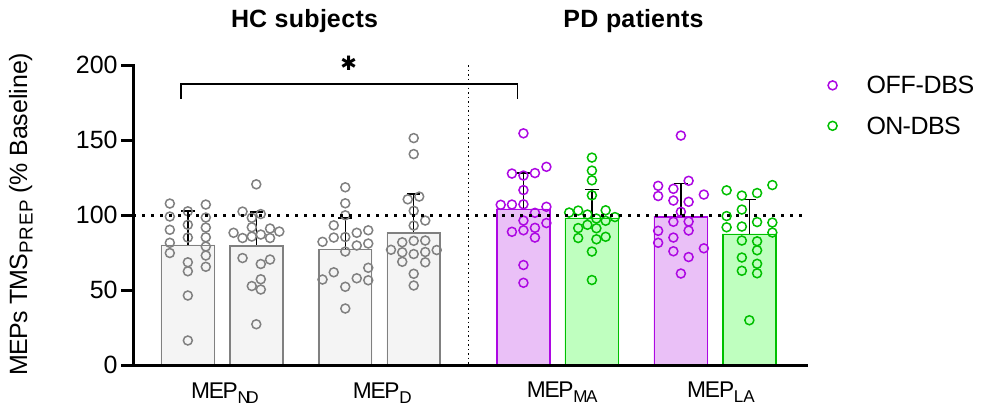

**Supplementary Figure 1. Preparatory suppression according to the DBS session,** in Parkinson’s disease (PD) patients (n=19), OFF-DBS (purple) and ON-DBS (green), and healthy control (HC) subjects (n=19) (all sessions in grey), after exclusion of outlier patient P8 (see Table 1 in the main manuscript) and the corresponding control subject. Note that here, for every participant, results of both RESP_SIDE_ conditions are pooled together. Each individual dot represents data from a single participant, while the bar plot represents the mean value (+ SD) for the condition. Neural measures of preparatory suppression are shown for the most-affected (MEP_MA_) and least-affected (MEP_LA_) hands. Taken together, this ﬁgure shows a tendency towards a more pronounced lack of preparatory suppression in the OFF- compared to the ON-DBS state in patients; post-hoc analyses, however, unveiled no significant difference between the two sessions, no matter the MEP_SIDE_ (both *P* > 0.38). Of interest is that MEPs acquired OFF-DBS on the MA side in patients were the only ones to signiﬁcantly differ from the corresponding condition in HC subjects. Mixed RM-ANOVA: **P* < 0.05.

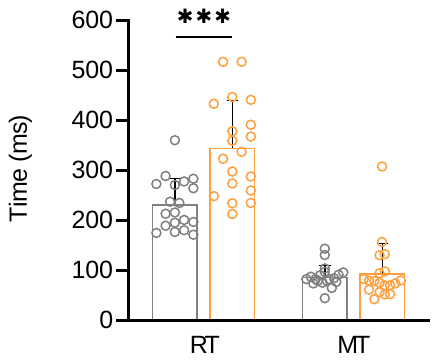

**Supplementary Figure 2. Reaction (RT) and movement time (MT) data (in ms)** in Parkinson’s disease (PD) patients (in orange) and healthy control (HC) subjects (in grey), after exclusion of outlier patient P8 (see Table 1, main manuscript) and the corresponding control subject. For every participant, results of both DBS sessions and both hands are pooled together, since the analysis did not reveal a signiﬁcant difference between sessions nor hands. Each individual dot represents data for a single subject, while the bar plot represents the mean value (+ SD) for the condition. RT were signiﬁcantly longer in patients compared to control subjects, while MT did not differ between groups, regardless of the disease or hand dominance. Mixed RM-ANOVA: ***P < 0.001.
